# Prospective Single Center Cohort Study for Early Diagnosis of Cardiac Dysfunction in Swiss Childhood Cancer Survivors: A Study Protocol

**DOI:** 10.1101/2020.01.10.20017152

**Authors:** Christina Schindera, Claudia E. Kuehni, Mladen Pavlovic, Eva Haegler-Laube, Nicolas Waespe, Jochen Roessler, Thomas M. Suter, Nicolas X. von der Weid, on behalf of the Swiss Pediatric Oncology Group (SPOG)

## Abstract

**Introduction:** Cardiovascular disease is the leading nonmalignant cause of late deaths in childhood cancer survivors. Cardiovascular disease and cardiac dysfunction can remain asymptomatic for many years, but eventually lead to progressive disease with high morbidity and mortality. Early detection and intervention are therefore crucial to improve outcome. In our study, we aim to 1) assess the prevalence of early cardiac dysfunction in adult childhood cancer survivors using conventional and speckle tracking echocardiography, 2) determine the association between cardiac dysfunction and treatment-related risk factors (anthracyclines, alkylating agents, steroids, cardiac radiation) and modifiable cardiovascular risk factors (abdominal obesity, hypertension), 3) investigate the development of cardiac dysfunction longitudinally in a defined cohort, 4) study the association between cardiac dysfunction and other health outcomes like pulmonary, endocrine, and renal diseases, quality of life, fatigue, strength and endurance, and physical activity, and 5) gain experience conducting a clinical study of childhood cancer survivors that will be extended to a national, multicenter study of cardiac complications.

**Methods:** We will invite ≥5 year childhood cancer survivors who were treated at the University Children’s Hospital Bern, Switzerland with any chemotherapy and/or cardiac radiation since 1976 and who are ≥18 years of age at time of study for a cardiac assessment at the University Hospital Bern. This includes overall 544 childhood cancer survivors, of whom about half were treated with anthracyclines and/or cardiac radiation and half with any other chemotherapy. The standardized cardiac assessment includes a medical history focusing on signs of cardiovascular disease and its risk factors, a physical examination, anthropometry, vital parameters, the 1-minute sit-to-stand test, and an echocardiography including speckle tracking.

**Results:** We will invite 544 eligible childhood cancer survivors for a cardiac assessment with a median age at study of 32.5 years and a median times since diagnosis of 25.0 years. Three hundred survivors (55%) are at high risk and 244 survivors (45%) are at standard risk for cardiac dysfunction.

**Discussion:** The results of this study will show the prevalence of early cardiac dysfunction in Swiss childhood cancer survivors, inform whether speckle tracking echocardiography is more sensitive to cardiac dysfunction than conventional echocardiography, and give a detailed picture of risk factors for cardiac dysfunction. The results will help improve primary treatment and follow-up care of children suffering from cancer.

**Trial registration:** Prospectively registered in ClinicalTrials.gov, Identifier: NCT03790943, registration date 25.06.2018.

## Introduction

Survival of childhood cancer has improved and the number of childhood cancer survivors (CCS) has greatly increased during recent decades [1, 2]. Consequently, more survivors face increased long-term morbidity and mortality due to chronic health conditions such as cardiovascular disease, pulmonary disease, and secondary neoplasms [3-5]. Among these, cardiovascular disease is the leading nonmalignant cause of death among CCS [3] with a cumulative incidence that increases up to 30 years after cancer diagnosis [6]. Heart failure, myocardial infarction, pericardial and valvular disease, and arrhythmias are all associated with treatments used in childhood cancer patients.

Studies from North America and The Netherlands assessed survivors exposed to cardiotoxic cancer therapy in whom prevalence of subclinical cardiac dysfunction from 6 to 27% was identified via conventional echocardiography [7, 8]. This suggests that many CCS have impaired cardiac function that might progress to clinical heart failure later in life. The North American study also found that a further 32% of survivors with otherwise normal conventional echocardiography showed evidence of cardiac dysfunction with abnormal strain measurements by speckle tracking echocardiography, a novel echocardiographic technique [7]. Additional studies have also suggested that speckle tracking echocardiography might be more sensitive to early cardiac dysfunction in childhood cancer survivors than conventional echocardiography [9, 10].

Most studies have assessed survivors exposed to anthracyclines and cardiac radiation, which are the most important treatment-related risk factors [11]. Yet other treatments may also increase childhood cancer survivors’ risk of cardiac dysfunction. Survivors who were not exposed to anthracyclines or cardiac radiation have demonstrated decreased left ventricular mass and increased cardiac biomarkers compared to siblings [12].

The North American Childhood Cancer Survivor Study analyzed self-reported data on cardiovascular risk factors in more than 10,000 adult CCS and showed that hypertension alone and in combination with other modifiable cardiovascular risk factors significantly increased the risk for heart failure, coronary artery disease, valvular disease, and arrhythmia in adult CCS [13]. The likelihood that modifiable cardiovascular risk factors might potentiate the increased risk of treatment-related cardiovascular disease in childhood cancer survivors thus motivates this study of cardiac dysfunction in adult CCSs.

## Methods

### Study objectives

The first and primary objective of this study is to assess the prevalence of early cardiac dysfunction in adult childhood cancer survivors using conventional and speckle tracking echocardiography. Second, we will determine the association between cardiac dysfunction and the treatment-related risk factors anthracyclines, alkylating agents, steroids, and cardiac radiation, and the modifiable cardiovascular risk factors abdominal obesity and hypertension. Our third objective is to investigate the development of cardiac dysfunction longitudinally in a defined cohort. Fourth, we will study the association between cardiac dysfunction and other health outcomes like pulmonary, endocrine, and renal diseases, quality of life, fatigue, strength and endurance, and physical activity. Finally, pursuing these objectives will provide experience conducting a clinical study of childhood cancer survivors that could be used for a national, multicenter study of cardiac complications.

### Primary outcome

The primary outcome of this study is echocardiographic parameters assessed by conventional and speckle tracking echocardiography (Table 1). Conventional echocardiography uses 2-dimensional (2D) and 3-dimensional (3D) left ventricular ejection fraction (LVEF) for systolic function, peak mitral flow velocity (E), early to late left ventricle filling velocity (E/A ratio), annular early diastolic velocity (e’), peak mitral flow velocity (E/e’ ratio), right atrium (RA) and right ventricle (RV), and the RV/RA ratio for diastolic function, valvular dysfunction, respiratory variation, and size of vena cava. Speckle tracking echocardiography includes longitudinal (LS), circumferential (CS), and radial strain (RS) for myocardial deformation imaging.

**Table 1.**
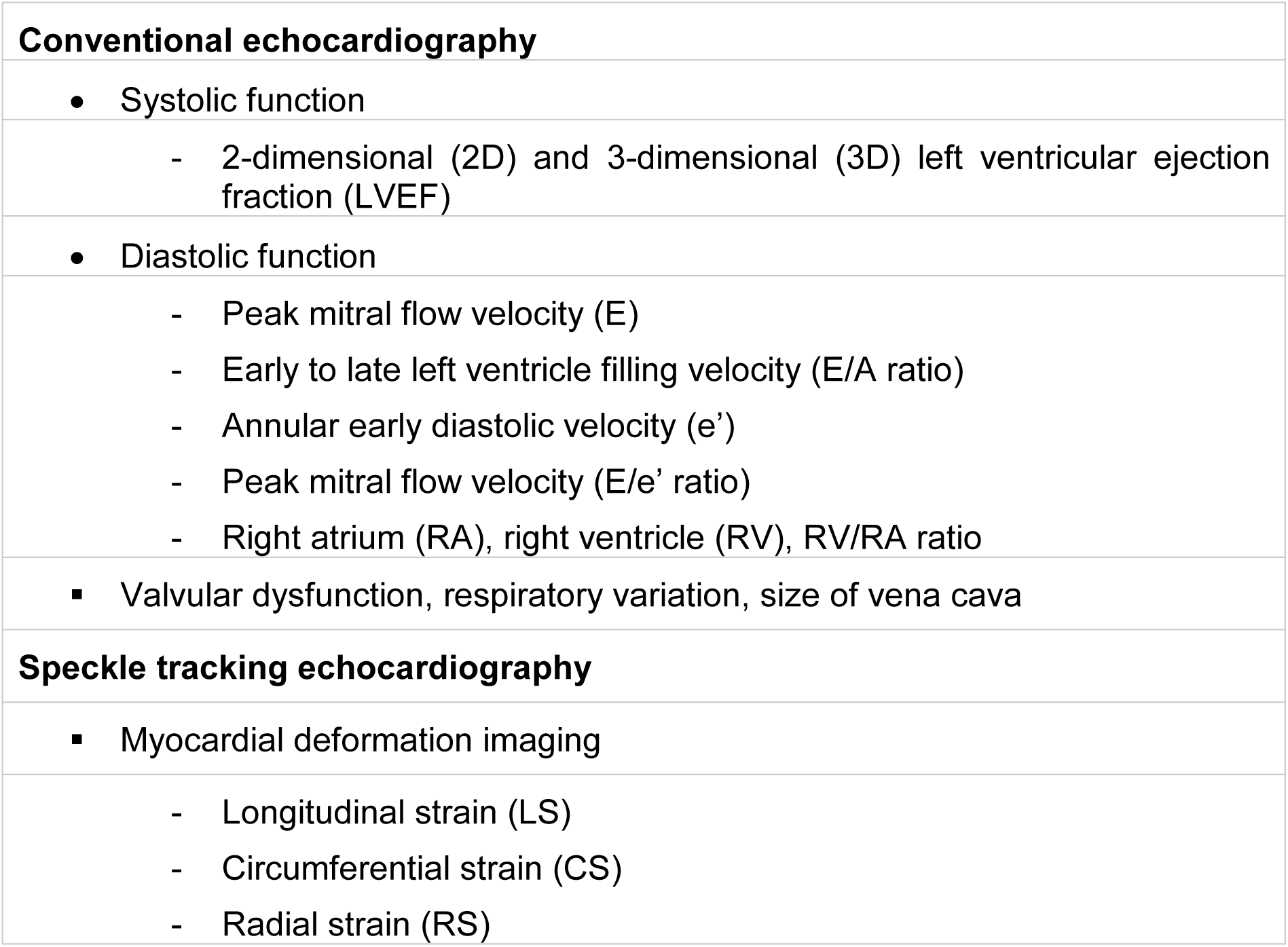
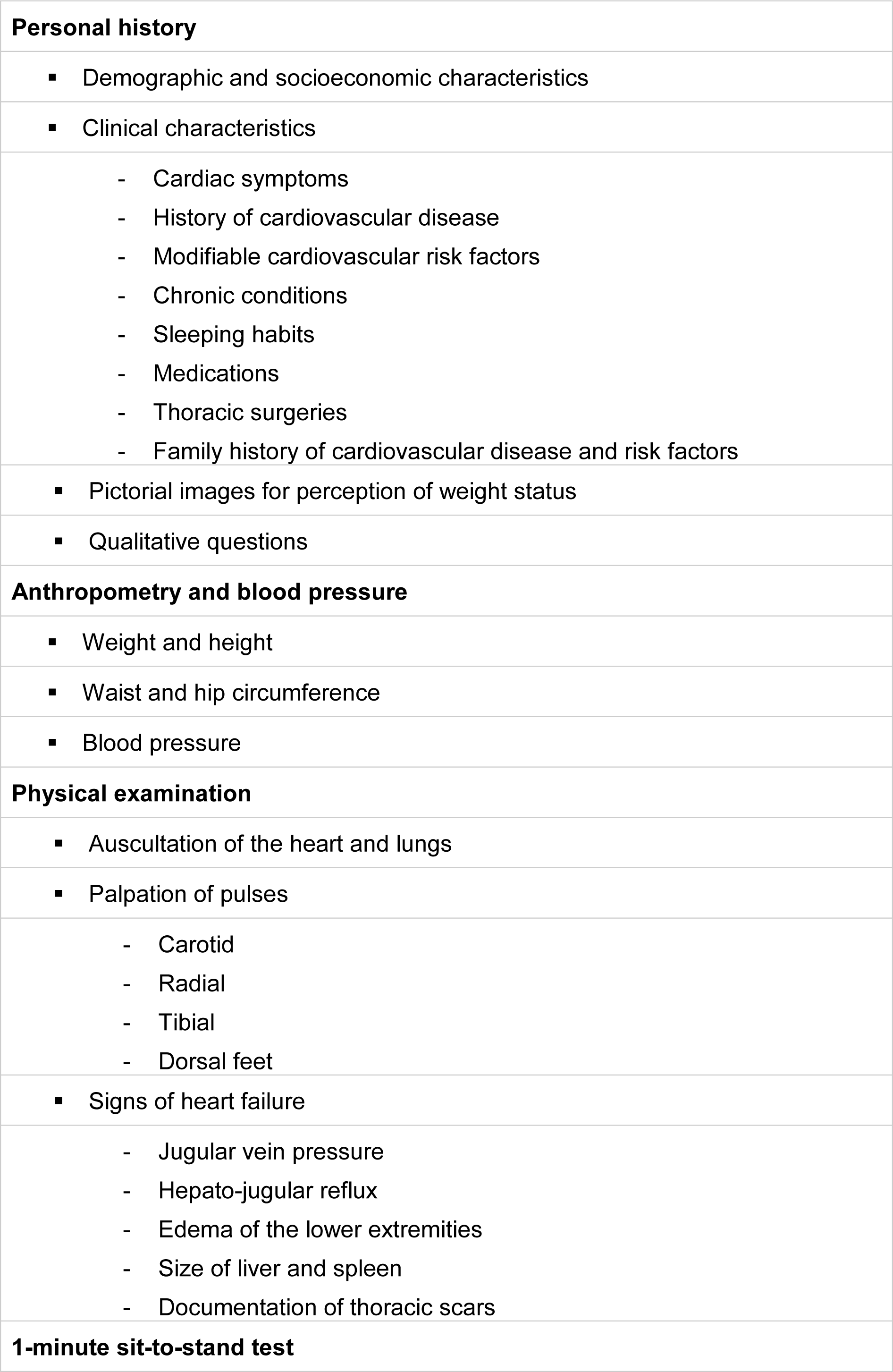
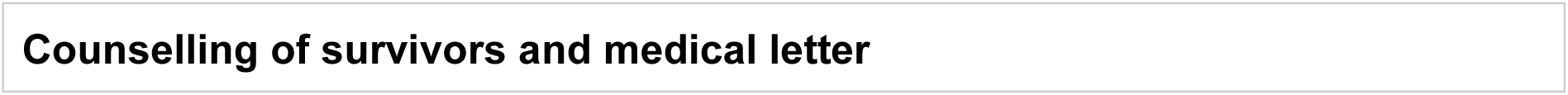
Components of cardiac assessment collected in childhood cancer survivors.

### Secondary outcomes

The secondary outcomes of this study assess the treatment-related risk factors anthracyclines, alkylating agents, steroids, and cardiac radiation; the modifiable cardiovascular risk factors abdominal obesity and hypertension; and other health outcomes including pulmonary, endocrine, and renal diseases, quality of life, fatigue, strength and endurance, and physical activity.

### Study design, study population, and inclusion criteria

This prospective cohort study is part of routine clinical follow-up care and is a collaborative and interdisciplinary effort of the Childhood Cancer Registry, the Swiss Childhood Cancer Survivor Study, the departments of Pediatric Hematology and Oncology, and Pediatric and Adult Cardiology at the University Hospital Bern, Switzerland (Table 2, Figure 1). The study includes all ≥5 year CCS diagnosed with childhood cancer since 1976 treated at the University Children’s Hospital Bern in Switzerland with any chemotherapy and/or cardiac radiation who are aged ≥18 years at study and registered in the Childhood Cancer Registry. The registry includes all patients in Switzerland diagnosed at age 0-20 years with leukemia, lymphoma, central nervous system tumors, malignant solid tumors, or Langerhans cell histiocytosis [14]. We classify cancer diagnoses according to the International Classification of Childhood Cancer, third edition (ICCC-3) into 12 main groups [15] and Langerhans cell histiocytosis. Recent estimates indicate that the registry includes >95% of children diagnosed below the age of 16 years since 1995 in Switzerland [16]. We exclude survivors who were treated with surgery or surgery and/or radiation other than cardiac radiation because these survivors have a standard risk of developing cardiac dysfunction. Ethics approval of this study is granted by the Ethics Committee of the Canton of Bern, Switzerland (KEK-BE: 2017-01612) and the study is registered at ClinicalTrials.gov (identifier: NCT03790943).

**Table 2.** Teams involved in the workflow of the study of early diagnosis of cardiac dysfunction

| Team | Involved staff |
| --- | --- |
| Childhood Cancer Registry | Administrative staff |
| Swiss Childhood Cancer Survivor Study | PhD student<br>Study nurse<br>Master students |
| Pediatric Hematology and Oncology | Head of pediatric hematology and oncology |
| Pediatric and Adult Cardiology | Cardiologists specialized in echocardiography<br>Cardiologist specialized in cardio-oncology<br>Nurse practitioner specialized in cardio-oncology<br>Administrative staff |

**Figure 1.**
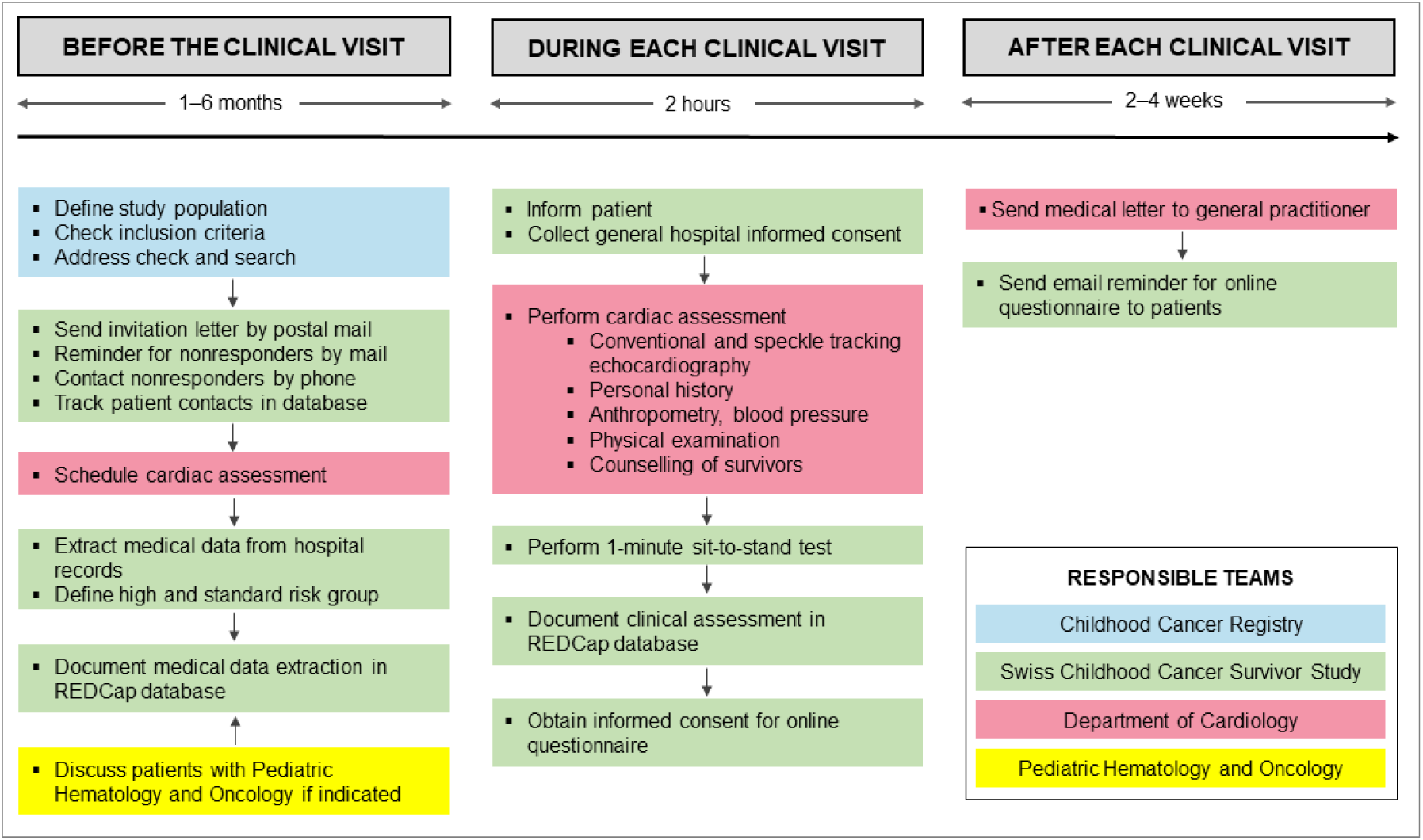
Responsible teams in the study of early diagnosis of cardiac dysfunction in childhood cancer survivors

### Study logistics

Current addresses of eligible survivors are obtained from the Childhood Cancer Registry and updated via the Swiss postal service where necessary (Figure 1). We send an invitation letter to survivors explaining why a cardiac assessment is useful, which describes planned examinations and the location of the visit in the Department of Cardiology at the University Hospital Bern. Survivors are asked to return a response form indicating their interest in participation in the study and the date and place of previous cardiac assessment(s). Nonresponders receive up to two reminders by mail before we try to contact them by phone. The administrative personnel of the Department of Cardiology schedules an appointment for a cardiac assessment via mail with survivors who agree to participate. The study is part of the routine follow-up care offered to CCS and is paid for by health insurance. All patient contacts are documented in a patient-tracking database.

### Medical data extraction

We extract the following data on each survivor from the cancer registry: cancer diagnose(s), relapse(s), age at cancer diagnosis, year of cancer diagnosis, and whether the person had chemotherapy, radiation (and if so the location of radiation), surgery, or hematopoietic stem cell transplantation (Figure 1).

We collect cumulative doses of anthracyclines, steroids, and alkylating agents from medical records (Table 3). We record the doses in each chemotherapy cycle together with patient weight and height. We calculate the cumulative doses per unit body-surface area expressed in milligrams per square meter. Cumulative anthracycline dose is calculated as doxorubicin-equivalent dose: [Doxorubicine x 1.0] + [Daunorubicine x 0.5] + [Idarubicine x 5] + [Epirubicine x 0.67] + [Mitoxandrone x 4] [17]. Cumulative dose of alkylating agents is calculated as cyclophosphamide-equivalent dose: [Cyclophosphamide x 1.0] + [Ifosfamide x 4.09] [18]. Cumulative steroid dose is calculated as prednisone-equivalent dose: [Prednisone x 1] + [Dexamethasone x 6.67] [19].

**Table 3.** Cumulative doses of chemotherapy and cardiac radiation extracted from medical records

| Cumulative doses of chemotherapy | Cumulative doses of cardiac radiation |
| --- | --- |
| Anthracyclines with doxorubicin-equivalent doses (mg/m<sup>2</sup>)<br>• Doxorubicin x 1.0<br>• Daunorubicin x 0.5<br>• Epirubicin x 0.67<br>• Idarubicin x 5.0<br>• Mitoxantron x 4.0 | Cardiac radiation (Gray)<br>• Chest<br>• Abdomen<br>• Whole or thoracic spine<br>• Total body irradiation |
| Alkylating agents with cyclophosphamide-equivalent doses (mg/m<sup>2</sup>)<br>• Cyclophosphamide x 1.0<br>• Ifosfamide x 4.09 |  |
| Steroids with prednisone-equivalent doses (g/m<sup>2</sup>)<br>• Prednisone x 1.0<br>• Dexamethasone x 6.67 |  |

Cardiac radiation includes radiation to the chest, abdomen, whole or thoracic spine, and total body radiation [17], and is collected from medical records (Table 3). We use the maximum documented dose of the field involving the heart and add the dose of total body irradiation.

### Definition of high risk and standard risk groups

Patients with exposure to any cumulative dose of anthracyclines and/or cardiac radiation (chest, abdomen, whole or thoracic spine, total body irradiation) are placed in the high risk group (Figure 2). High risk patients are assessed longitudinally with baseline and follow-up cardiac assessments according to the Children’s Oncology Group Long-Term Follow-Up Guidelines, Version 5.0, October 2018 [17]. Survivors with exposure to any chemotherapy other than anthracyclines are assigned to a standard risk group and evaluated cross-sectionally unless a cardiac follow-up assessment is clinically indicated. All survivors with only surgery or radiation other than cardiac are excluded from this study and are seen only within the routine follow-up care without echocardiography.

**Figure 2.**
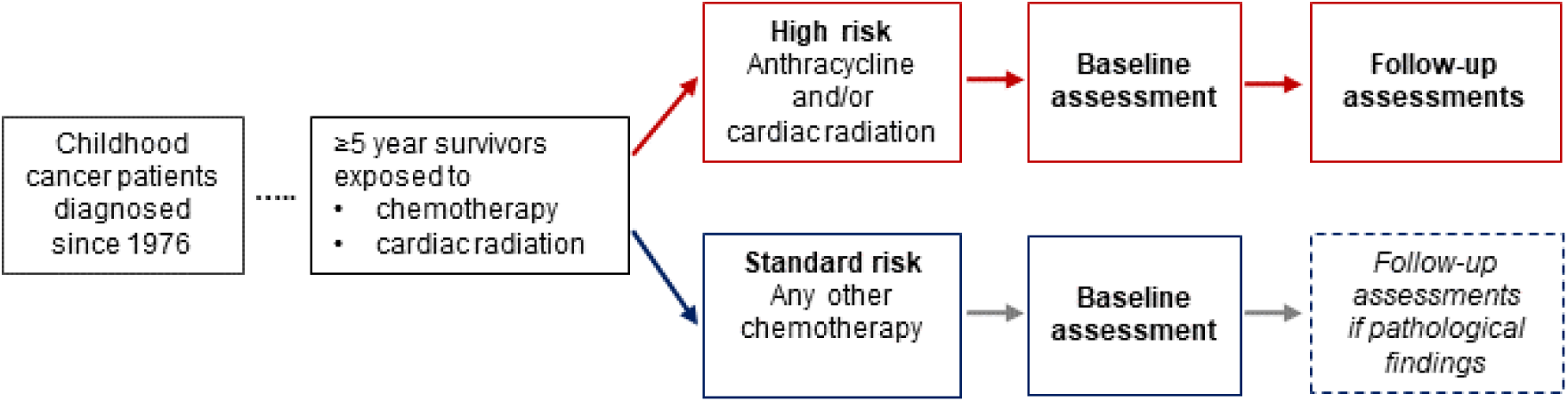
Study design and risk group stratification of childhood cancer survivors.

### Patient information and informed consent

At the cardiac assessment, we give survivors oral and written information about the clinical study (Figure 1). The general hospital informed consent form that participants sign is a standard consent form widely used in Swiss inpatient and outpatient settings to enable research with clinical data.

### Echocardiography

Echocardiography is performed by experienced cardiologists from the Department of Cardiology who are blinded with respect to the patient’s cancer treatment and risk group (Table 2). Conventional echocardiography includes assessment of systolic function (2D and 3D LVEF), diastolic function (E, E/A ratio, e’, E/e’ ratio, RA, RV, RV/RA ratio), valvular dysfunction, respiratory variation, and size of vena cava (Table 1, Figure 1) using a GE E9 or E95 (GE Vingmed, Norway). Speckle tracking echocardiography includes LS, CS, and RS and is performed using EchoPAC PC version 112 (GE Vingmed, Norway).

### Personal history

We take a comprehensive personal history of survivors’ demographic and socioeconomic characteristics: current occupation, employment status, work hours per week, marital status, offspring, and housing situation (Table 1, Figure 1). We also obtain clinical characteristics including cardiac symptoms (New York Heart Association I-IV), history of cardiovascular disease, modifiable cardiovascular risk factors (smoking, physical activity, drug consumption), chronic conditions (pulmonary, endocrine, and renal diseases), sleeping habits, medications, thoracic surgeries, and family history of cardiovascular diseases and risk factors. To determine perception of weight status we use pictorial images of women and men after Harris et al. [20] and ask the patient to indicate the picture that best matches the weight status of his/her parents, siblings, and the patient’s own weight. We also ask patients, Was it a big effort for you to come to the hospital for today’s appointment, does your history of childhood cancer play a role in your daily life, and are you afraid that your treatment for cancer during childhood caused any medical problems in adulthood? We also ask patients if they have any questions, requests, or wishes to direct to us.

### Anthropometry and blood pressure

Weight and standing height are taken by standard procedures, barefoot and in light clothes (Table 1, Figure 1). Weight is determined to the nearest 0.1 kg and height to the nearest 0.5 cm. Body mass index (BMI) is expressed as kg/m^2^ [21]. Waist and hip circumferences are measured using a measuring tape to the nearest 0.1 cm. Waist circumference is measured at the midpoint between the lower margin of the lowest rib and the top of the iliac crest and hip circumference at the widest circumference over the buttocks [21]. The waist-hip ratio is calculated as waist circumference divided by hip circumference. Blood pressure is measured comfortably in a quiet environment with three measurements repeated in a sitting position and the average of the last two readings is recorded. Additional measurements are taken if the first two readings of systolic or diastolic blood pressure differ by >10 mm Hg [22].

### Physical examination

We perform a thorough physical examination with special emphasis on signs of cardiovascular disease (Table 1, Figure 1). This includes auscultation of the heart and lungs, palpation of the carotid, radial, tibial, and dorsal foot artery pulses, examination of the jugular vein pressure, hepato-jugular reflux, edema of the lower extremities, size of liver and spleen, and documentation of thoracic scars.

#### 1-minute sit-to-stand test

We perform the 1-minute sit-to-stand test (STS) which captures the number of times a person can stand up and sit down on a regular chair in one minute (Table 1, Figure 1) [23]. The STS is an estimate of lower body muscular strength and endurance. We compare our population with population-based, age- and sex-adjusted Swiss reference values [23].

### Counselling of survivors and medical letter

At the end of the cardiac assessment, a cardiologist specialized in cardio-oncology and who did not perform the echocardiography explains the results of the echocardiography to survivors and counsels them on their cardiac function and the presence (or absence) of modifiable cardiovascular risk factors (Table 1, Figure 1). Recommendations on follow-up assessments are based on the Children’s Oncology Group Long-Term Follow-Up Guidelines, Version 5.0, October 2018 [17]. A medical letter summing up the results of the cardiac assessment is sent to the survivor’s general practitioner.

### Online questionnaire

We ask survivors to answer four questionnaires after returning home. The Short Form 36 Health Survey (SF-36) assesses health-related quality of life [24] and has been used before in CCS [25]. The Seven-Day Physical Activity Recall questionnaire (PAR) measures moderate and vigorous physical activity, and sleep during the last seven days [26]. Fatigue is assessed using the Checklist Individual Strength (CIS), a validated 20-item questionnaire that identifies different aspects of fatigue within the previous two weeks [27]. Diet and alcohol consumption are obtained using questions from the SCCSS questionnaire [28]. We ask survivors to give separate informed consent for the online questionnaires. Survivors not completing the online questionnaires within two weeks after the clinical visit are reminded by email.

## Documentation

All parts of the cardiac assessment are directly entered into a dedicated REDCap (version 8.5.19, Vanderbilt University, Nashville, TN, USA) database to minimize risk of disclosure. Within the database, each survivor has a unique identification number (ID). No personal information can be obtained with this number. Data containing survivors’ unique IDs are stored on encrypted devices or secured servers at the University of Bern.

### Cardiac assessment, phase 1

In 2016-2017, we started a cardiac assessment using standardized echocardiography as described (Figure 3). At that time, personal history, blood pressure, physical examination, and the STS were not yet assessed in a standardized way. The personal history and the online questionnaires were retrospectively completed by phone interviews between 05/2017–06/2017.

**Figure 3.**
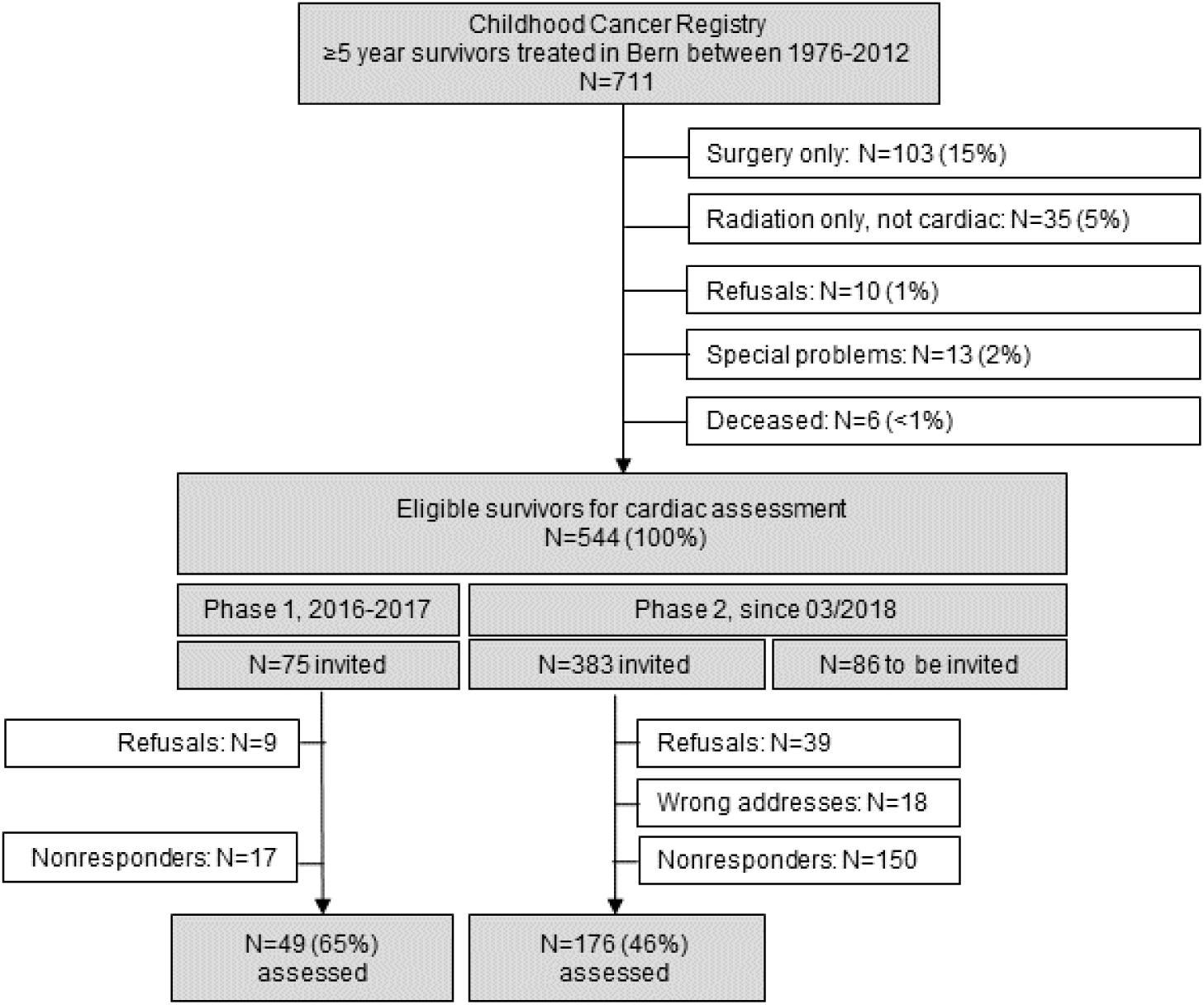
Population tree of childhood cancer survivors eligible for the study, status by October 1^st^, 2019

### Cardiac assessment, phase 2

Since March 2018, we have been collecting all data in a standardized way. Data collection is performed by the study team from the Childhood Cancer Registry, the Swiss Childhood Cancer Survivor Study, the Department of Pediatric Hematology and Oncology, and the Department of Cardiology at the University Hospital Bern (Table 2, Figure 3).

### Statistical analyses

We will compare characteristics of responders and nonresponders using chi-square tests and perform univariable and multivariable logistic regression to investigate the association between cardiotoxic treatment exposures (anthracyclines, alkylating agents, steroids, cardiac radiation) and modifiable cardiovascular risk factors (abdominal obesity, hypertension), and cardiac dysfunction adjusting for sex, age at study, and follow-up time. We use STATA software (Version 15.1, Stata Corporation, Austin, TX, USA).

## Results

### Patient population

On January 1, 2018, the Childhood Cancer Registry included 711 survivors aged ≥18 years who had been diagnosed and treated at the University Children’s Hospital Bern since 1976 and had survived ≥5 years (Figure 3). Among those, 103 were excluded because of surgery as only treatment, 35 survivors because of radiation only other than cardiac, and 29 survivors because they did not want to be contacted or who had specific problems (e.g. did not want to be invited for clinical studies because of emotional stress), and because they had died. Among these survivors, 544 met the inclusion criteria for an invitation to a clinical visit (Table 4, Figure 3). This number includes 300 survivors (55%) at high risk for cardiac dysfunction and 244 survivors (45%) at standard risk for cardiac dysfunction with a median age at study of 32.5 years and a median times since diagnosis of 25.0 years (Table 4). In Phase 1 (2016-2017), 75 survivors were invited and 49 survivors attended the cardiac assessment with a response rate of 65%; phase 2 is ongoing (Figure 3). We plan to recruit new five-year survivors continuously into the study, so the size of the cohort will increase and numbers will change.

**Table 4.**
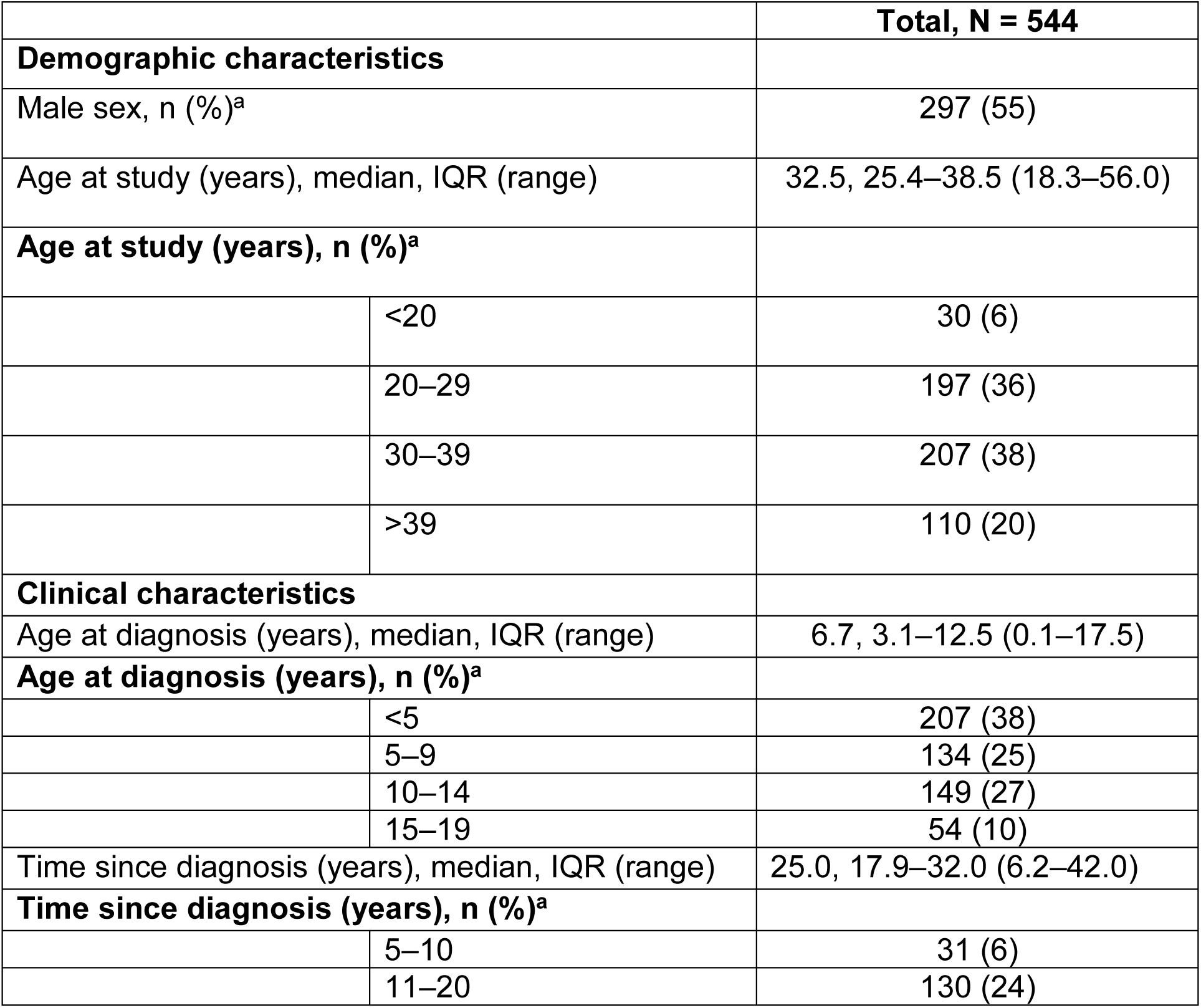
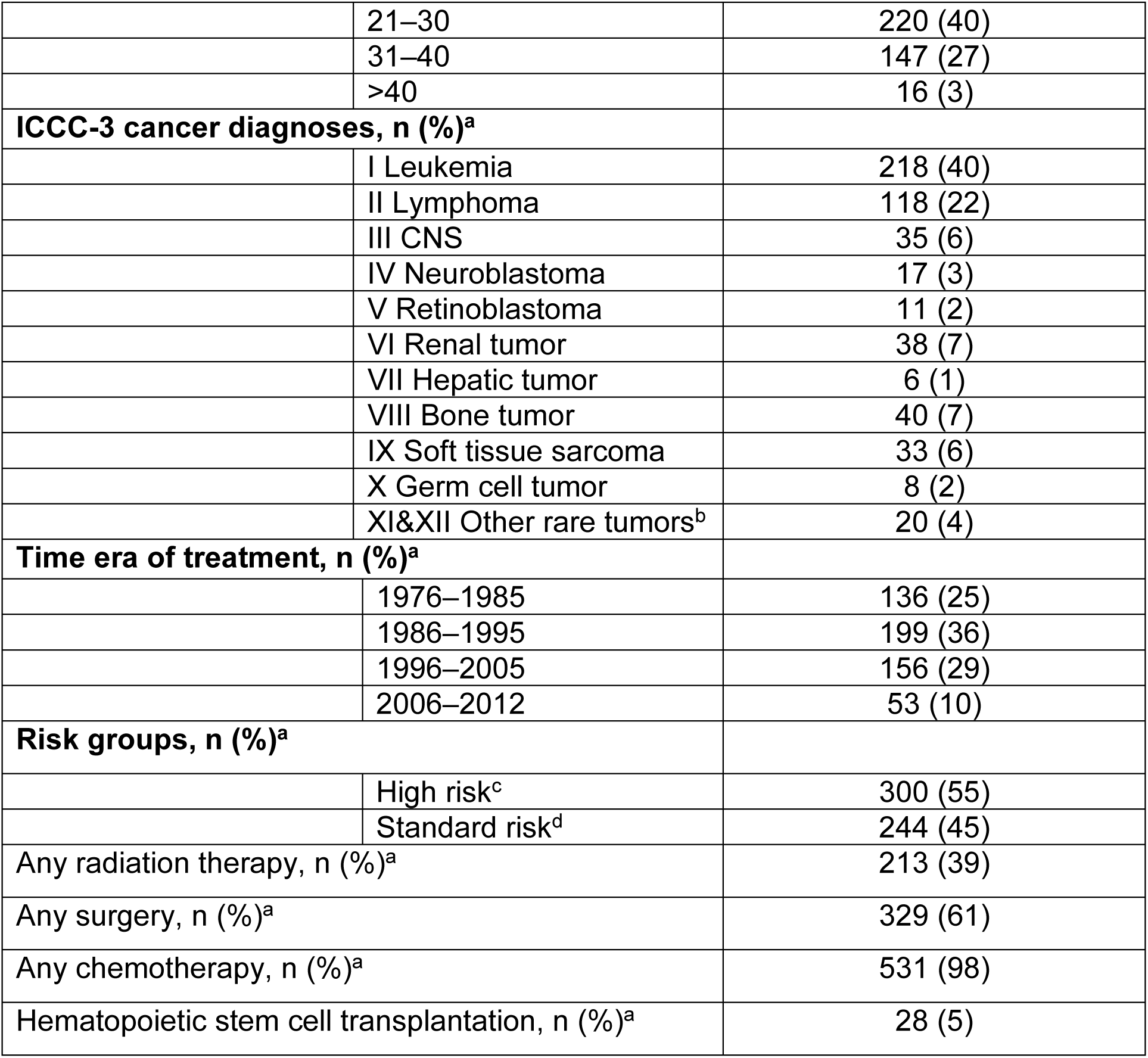
Demographic and clinical characteristics of survivors eligible for participation in the cohort study

## Discussion

This prospective, single-center cohort study investigates the prevalence of cardiac dysfunction and its risk factors in adult childhood cancer survivors, and compares conventional and speckle tracking echocardiography.

Few studies are comparable to ours. A single-center study in The Netherlands included 525 adult CCS who had been treated between 1966–1997 with anthracyclines, high-dose cyclophosphamide, high-dose ifosfamide, and/or cardiac radiation [8]. Conventional echocardiography was performed between 1996–2004 to measure left ventricular shortening fraction and subclinical cardiac dysfunction was observed in 27% of survivors at a median follow-up time of 15 years. Another hospital-based, single-center study at St. Jude, USA, assessed 1820 adult CCS exposed to anthracyclines and/or cardiac radiation at a median follow-up time of 23 years using conventional and speckle tracking echocardiography [7]. One-third of survivors with normal LVEF had abnormal longitudinal strain by speckle tracking echocardiography. Risk factors for pathological findings in conventional and speckle tracking echocardiography were anthracyclines and cardiac radiation. The modifiable cardiovascular risk factors hypertension, abdominal obesity, dyslipidemia, and high fasting glucose were associated with abnormal longitudinal strain but not with LVEF, suggesting that speckle tracking might be more sensitive in detecting cardiac dysfunction.

First among this study’s limitations is that currently it is confined to a single center. However, the nine centers treating children and adolescents with cancer in Switzerland collaborate closely and use uniform treatment protocols; we therefore expect that results from the University Hospital Bern are representative of all nine centers in the country. We also are concerned that our study includes a heterogeneous group of CCS with relatively small numbers in each subgroup defined by treatment exposure or type of cancer. We plan to overcome this limitation by expanding this study to a nationwide study that includes all nine Swiss Pediatric Oncology Group clinics. Finally we note that echocardiographic procedures can be somewhat investigator dependent. We try to overcome this by strict adherence to our standard operating procedures. Furthermore, the advantage of strain analysis is a smaller inter-observer variability with strain analysis compared to LVEF-measurements in 2D echocardiography [29]. Intra-observer as well as inter-observer reproducibility is even better in 3D echocardiography compared with conventional 2D echocardiography if imaging quality is high [30]. By using only one type of vendor equipment and software we furthermore avoid inter-vendor variability.

Among this study’s several strengths is our attempt to include the complete cohort of survivors treated at the University Children’s Hospital Bern since 1976 based on the database of the Childhood Cancer Registry. We repeat our invitation to nonresponders several times and ask about reasons for not participating. This reduces the potential for selection bias. Second, we have access to all treatment exposures based on actual chemotherapy road maps and are able to look into dose-response relationships. Third, we will continuously include new five-year survivors and will therefore gain knowledge about the risk for cardiac dysfunction in younger patients treated more recently. Finally, our study has been set up within routine survivorship follow-up care using the experience of a multi- and interdisciplinary team with close collaboration between pediatric and adult cardiology, pediatric hematology and oncology, and clinical epidemiology.

The preliminary results from this prospective, single-center study suggest that a standardized cardiac assessment that is part of routine follow-up care done in collaboration between pediatric and adult specialists is feasible in Switzerland and widely accepted by survivors and health care providers. In the next step, we will include more Swiss centers in the study to provide standardized clinical follow-up care longitudinally to all childhood cancer survivors on a nationwide scale.

## Data Availability

Researchers interested in collaborative work can contact the corresponding author (Claudia E. Kuehni) to discuss planned projects or analyses of existing data. The final decision will be made upon presentation of the project to the study’s principal investigators.

## List of abbreviations

2D: 2-dimensional
3D: 3-dimensional
BMI: Body mass index
CCS: Childhood cancer survivors
CIS: Checklist Individual Strength
CS: Circumferential strain
E: Peak mitral flow velocity
E/A: Early to late left ventricle filling velocity
e’: Annular early diastolic velocity
E/e’: Peak mitral flow velocity
ICCC3: International Classification of Childhood Cancer third edition
IQR: Interquartile range
LS: Longitudinal strain
LVEF: Left ventricular ejection fraction
N: Numbers
PAR: Seven-Day Physical Activity Recall questionnaire
RA: Right atrium
RS: Radial strain
RV: Right ventricle
SF-36: Short Form 36 Health Survey
STS: 1-minute sit-to-stand test

## Declarations

### Ethics approval and consent to participate

The study was approved by the Swiss Ethics Committees on research involving humans (Kantonale Ethikkomission Bern [KEK]; reference number: KEK-BE: 2017-01612). Informed consent as documented by signature is obtained from each survivor prior to participation in the study.

### Consent for publication

Not applicable as this manuscript does not contain data from individual survivors.

### Competing interests

The authors declare that they have no competing interests.

### Funding

This study is supported by the Swiss Cancer League (KLS-3886-02-2016) which funds the salary of a PhD-student (CS) and a study nurse. The Stiftung für krebskranke Kinder, Regio basiliensis (AND-4641-01-2018) gave a travel grant to CS for a research visit to St. Jude Children’s Research Hospital, Memphis, TN, USA. The work of the Childhood Cancer Registry is supported by the Swiss Pediatric Oncology Group (www.spog.ch), Schweizerische Konferenz der kantonalen Gesundheitsdirektorinnen und -direktoren (www.gdk-cds.ch), Swiss Cancer Research (www.krebsforschung.ch), Kinderkrebshilfe Schweiz (www.kinderkrebshilfe.ch), the Federal Office of Public Health (FOPH), and the National Institute of Cancer Epidemiology and Registration (www.nicer.org).

### Author’s contributions

NvdW, MP, TS, CEK, and CS have designed the study and developed the study material. MP, TS, CEK, NvdW, CS, EHL, and JR participated in the study management and coordination. CS, CK, and NvdW drafted the manuscript. All authors commented and approved the final version.

## Acknowledgements

We thank all childhood cancer survivors for participating in our study. We thank Daniel Rhyner and Michele Martinelli for performing the clinical assessment. We thank Susanne Suter, Nadine Lötscher, Caleb Leung, Pascale Annaheim, and Annina Elmiger for supporting the study and giving valuable inputs. We thank the study team of the SCCSS: Fabiën Belle, Carole Dupont, Rahel Kasteler, Rahel Kuonen, Jana Remlinger, Grit Sommer, Maria Otth, and Annette Weiss. We also thank the data managers of the SPOG: Dr. Claudia Althaus, Nadine Assbichler, Pamela Balestra, Heike Baumeler, Nadine Beusch, Sarah Blanc, Dr. Pierluigi Brazzola, Susann Drerup, Janine Garibay, Franziska Hochreutener, Monika Imbach, Friedgard Julmy, Eléna Lemmel, Rodolfo Lo Piccolo, Heike Markiewicz, Dr. Veneranda Mattielo, Annette Reinberg, Dr. Renate Siegenthaler, Astrid Schiltknecht, Beate Schwenke, and Verena Stahel. And we thank the SCCR team of Meltem Altun, Erika Brantschen, Katharina Flandera, Elisabeth Kiraly, Verena Pfeiffer, Shelagh Redmond, Julia Ruppel, Ursina Roder. For editorial assistance we thank Christopher Ritter.

## Notes

### Competing Interest Statement

The authors have declared no competing interest.

### Clinical Trial

ClinicalTrials.gov, Identifier: NCT03790943

